# A pattern categorization of CT findings to predict outcome of COVID-19 pneumonia

**DOI:** 10.1101/2020.05.19.20107409

**Authors:** Chao Jin, Yan Wang, Carol C. Wu, Huifang Zhao, Ting Liang, Zhe Liu, Zhijie Jian, Runqing Li, Zekun Wang, Fen Li, Jie Zhou, Shubo Cai, Yang Liu, Hao Li, Zhongyi Li, Yukun Liang, Heping Zhou, Xibin Wang, Zhuanqin Ren, Jian Yang

**Author notes:** **Corresponding author**: Jian Yang, Ph.D., M.D. Professor, Department of Radiology, The First Affiliated Hospital of Xi’an Jiaotong University, Xi’an 710061, PR China.

## Abstract

**Purpose:** As global healthcare system is overwhelmed by novel coronavirus disease (COVID-19), early identification of risks of adverse outcomes becomes the key to optimize management and improve survival. This study aimed to provide a CT-based pattern categorization to predict outcome of COVID-19 pneumonia.

**Methods:** 165 patients with COVID-19 (91 men, 4-89 years) underwent chest CT were retrospectively enrolled. CT findings were categorized as Pattern0 (negative), Pattern1 (bronchopneumonia), Pattern2 (organizing pneumonia), Pattern3 (progressive organizing pneumonia) and Pattern4 (diffuse alveolar damage). Clinical findings were compared across different categories. Time-dependent progression of CT patterns and correlations with clinical outcomes, i.e. discharge or adverse outcome (admission to ICU, requiring mechanical ventilation, or death), with pulmonary sequelae (complete absorption or residuals) on CT after discharge were analyzed.

**Results:** Of 94 patients with outcome, 81(86.2%) were discharged, 3(3.2%) were admitted to ICU, 4(4.3%) required mechanical ventilation, 6(6.4%) died. 31(38.3%) had complete absorption at median day 37 after symptom-onset. Significant differences between pattern-categories were found in age, disease-severity, comorbidity and laboratory results (all *P*<0.05). Remarkable evolution was observed in Pattern0-2 and Pattern3-4 within 3 and 2 weeks after symptom-onset, respectively; most of patterns remained thereafter. After controlling for age, CT pattern significantly correlated with adverse outcomes (Pattern4 vs. Pattern0-3 [reference]; hazard-ratio[95%CI], 18.90[1.91-186.60], *P*=0.012). CT pattern (Pattern3-4 vs. Pattern0-2 [reference]; 0.26[0.08-0.88], *P*=0.030) and C-reactive protein (>10 vs. ≤10mg/L [reference]; 0.31[0.13-0.72], *P*=0.006) were risk-factors associated with pulmonary residuals.

**Conclusion:** CT pattern categorization allied with clinical characteristics within 2 weeks after symptom-onset would facilitate early prognostic stratification in COVID-19 pneumonia.

## Introduction

Since the latter part of December of 2019, an outbreak of respiratory disease caused by severe acute respiratory syndrome-coronavirus-2 (SARS-CoV-2) has become a pandemic [1]. As of May 4, 2020, 3,356,205 laboratory-confirmed cases and 238,730 deaths have been reported [2]. Numerous studies have revealed the epidemiological, clinical and radiological characteristics of the novel coronavirus disease (COVID-19) [3-6]. Despite the fact that more than 80% of infected patients manifest with only mild clinical symptoms^3^, early identifying the risks of an adverse outcome remains the key to optimize management and improve survival. Previous studies found that advanced age and presence of comorbidity (e.g. cardiovascular disease or hypertension) were risk factors associated with an adverse outcome such as admission to intensive care unit (ICU), need for mechanical ventilation, or death [7,8]. In addition, some laboratory indicators e.g. elevated hypersensitive troponin I, leukocytosis, neutrophilia, lymphopenia and elevated D-dimer were found to be linked with unfavorable clinical outcomes [7-9]. Presence of consolidation on computed tomography (CT) was also considered to be predictive of poor outcome in COVID-19 [10]. Despite the above, the identification of early prognostic signs of COVID-19 remains of urgent importance due to the diversity in clinical and imaging findings as well as the severity and rapid progression of disease.

It is recognized that CT plays a central role in diagnosis and management of COVID-19 pneumonia [11-13]. Reported CT findings of COVID-19 pneumonia included the ground glass opacities (GGO), consolidation, septal thickening mainly along the subpleural lungs or bronchovascular bundles or diffusely in the entire lungs [14]. These are highly suggestive of lung organization response to injury from COVID-19 pneumonia, similar to radiological findings in the diffuse alveolar damage (DAD) and organizing pneumonia (OP) [15]. Pathological studies also observed DAD in patients who succumbed to COVID-19 [16]. Previous studies have demonstrated a decreased survival rate of 35-50% in DAD, while most patients with OP had better prognosis [15]. In this regard, a pattern categorization of COVID-19 pneumonia, i.e. DAD and OP patterns may help the prognostic stratification. Based on the prior study regarding influenza A (H1N1) pneumonia [17], Lee also suggested a pattern categorization of COVID-19, i.e. bronchopneumonia, OP and DAD [18]. A rapid progression of OP-like injury in Severe Acute Respiratory Syndrome (SARS) was considered to be predictive of a protracted clinical course [19]. This may suggest a progressive subtype of OP pattern. Based on the aforementioned knowledge, a CT pattern categorization of COVID-19 pneumonia, i.e. bronchopneumonia, OP, progressive OP and DAD may have potential prognostic implications, e.g. adverse outcome, clinical course with recovery. As healthcare systems in many countries are overwhelmed with COVID-19 patients, improved prediction of the course of the disease based on early findings can assist with improved utilization of limited resources. To this end, this study aimed to investigate the prognostic significance of a CT pattern categorization in conjunction with the clinical indicators on clinical outcome and pulmonary sequelae in COVID-19.

## Methods

### Participants

The internal review board approved this retrospective study. Written informed consent was waived with approval. Between January 22, and March 16, 2020, 172 laboratory-confirmed COVID-19 patients who underwent chest CT were collected from 8 hospitals in China. The cases were from 4 regions (Xi’an, *n*=80; Baoji, *n*=10; Ankang, *n*=18; Hanzhong, *n*=17) in Shaanxi province and Wuhan (*n*=47) in Hubei province.

A case of COVID-19 was confirmed by a positive result on next-generation sequencing or real-time RT-PCR. The disease type, i.e. uncomplicated illness, mild pneumonia, severe pneumonia, critical illness (acute respiratory distress syndrome, sepsis or septic shock) was evaluated based on the criteria published by World Health Organization (WHO) [20].

All the patients were treated based on Diagnosis and Treatment of Pneumonia Caused by COVID-19 (version 7) issued by National Health Commission of the People’s Republic of China1, which includes initiation of antivirals, interferon, Chinese herbal medications, supplemental oxygen as needed and hospitalization. The criteria for patient discharge with recovery included: (1) afebrile for >3 days, (2) improved respiratory symptoms, (3) chest imaging shows obvious resolution of inflammation, and (4) two consecutively negative nucleic acid test results (sampling interval ≥ 1 day) [21]. The recommendations for discharged patients included (1) 14 days of isolation management and health monitoring; (2) follow-up hospital visits with a next-generation sequencing or real-time RT-PCR test and chest CT scan to detect whether there exist a positive return and/or pulmonary residuals excluding the underlying lesions on CT with linear opacities, and/or a few consolidation with/without GGO at 2 and 4 weeks after discharge [21].

### CT image acquisition

All chest CT were acquired by using 16- or 64-multidector CT scanners (GE LightSpeed 16, GE VCT LightSpeed 64, GE Optima 680, GE Healthcare; Philips Brilliant 16, Philips Healthcare; Somatom Sensation 64, Somatom AS, Somatom Spirit, Siemens Healthcare). Patients were scanned in the supine position from the level of the upper thoracic inlet to the inferior level of the costophrenic angle with the following parameters: tube voltage of 120 kVp, current intelligent control (auto mA) of 30-300 mA, and slice thickness reconstructions of 0.6-1.5 mm.

### Data collection and evaluation

We extracted the demographic data, clinical symptoms, and laboratory tests on admission from electronic medical records. The date of disease onset was defined as patients’ reported date of symptom onset. The time intervals from symptom onset to each CT were determined. The primary clinical outcome was discharge or adverse outcome (admission to ICU, use of mechanical ventilation, or death). The secondary outcome was pulmonary sequelae, i.e. complete absorption or residuals on CT at the first follow-up visit after discharge.

All CT images and pattern categorization were independently evaluated by two experienced radiologists respectively with 4 and 10 years of pulmonary imaging experience, who were blinded to the clinical and laboratory data of patients. Prior to the evaluation, they were trained by a lecture- and literature-based session that explained CT findings [10-13], a chest imaging score assessing the degree of lobar involvement [22], and pattern categorizations [15,17] of COVID-19. During the session, 209 CT images from 56 cases randomly selected from this study cohort were individually evaluated and then differences were discussed with a final consensus. The remaining CT images were first individually evaluated and then evaluated together 3 weeks after individual evaluation. Any difference was discussed with a final consensus. Individual evaluations were used for calculation of inter-observer agreement (See more in the Supplement), and consensus evaluations were used for subsequent analysis.

CT findings including the presence and distribution of GGO, consolidation, linear opacity, pleural effusion and lymphadenopathy were evaluated. The degree of lobar involvement and total lung severity score were also evaluated [22]. Based on the degree or area of involvement, each of the five lung lobes was scored of 0 for 0 % lobe involvement, 1 for 1-25% lobe involvement, 2 for 26-50% lobe involvement, 3 for 51-75% lobe involvement, or 4 for 76-100% lobe involvement. A total severity score was calculated by summing the scores of the five lobes (range, 0-20).

CT pattern categorization was performed by taking previous reports [15,17] (Table 1). In cases with two or more patterns, predominant pattern was designated.

**Table 1.** Definition of COVID-19 pneumonic pattern based on CT findings.

| CT pattern | Definition | CT findings |
| --- | --- | --- |
| Pattern 0 | Negative | None |
| Pattern 1 | Bronchopneumonia pattern | Discrete lesion with a peribronchial distribution<br>CT signs with GGO or consolidation, or tree-in-bud sign or nodular opacity (Figure 3)<br>Lung lobar involvement assessed by total CT score less than or equal to 5 |
| Pattern 2 | Organizing pneumonia pattern | Multifocal lesions with a peripheral distribution predominantly in the middle to lower lung zones<br>CT signs with GGO or consolidation, and/or interlobular septal thickening (Figure 4)<br>Lung lobar involvement assessed by total CT score less than or equal to 6 |
| Pattern 3 | Progressive organizing pneumonia pattern | Multiple lesions with a peripheral distribution predominantly in the middle to lower lung zones<br>CT signs with consolidation or GGO or mixed GGO and consolidation, and/or interlobular septal thickening (Figure 5)<br>Lung lobar involvement assessed by total CT score more than 6 and less than 10 |
| Pattern 4 | Diffuse alveolar damage pattern | Lesions with extensive distribution diffusely in the entire lungs<br>CT signs with consolidation mixed with or without GGO, and/or air bronchograms (Figure 6)<br>Lung lobar involvement assessed by total CT score more than or equal to 10 |

Note: The primary CT signs (GGO, consolidation, linear opacity, interlobular septal thickening and air bronchograms) were included to define the CT patterns; while other signs e.g. pleural effusion, lymphadenopathy and so on were not considered due to the infrequency in each pattern<sup>12</sup>.
Abbreviations: GGO = ground glass opacities.

### Statistical Analysis

Continuous variables were represented as means and standard deviations, while categorical variables were expressed as counts and percentages. Differences of demographic, clinical and CT imaging characteristics across pattern groups were analyzed by dependent sample t-test, Chi-square test or Fisher’s exact test as appropriate. Bonferroni correction was used in multiple comparisons. Chi-square test for trend was used to explore the time-dependent change of each CT pattern. Univariate Cox proportional-hazards regression was first used to explore the risk factors related to clinical adverse outcomes and pulmonary residuals. Multivariate Cox proportional-hazards regression with Kaplan-Meier curve plots were further used to explore the risk factors based on the significant variables in the above univariate analysis.

All statistical analyses were performed using SPSS 17.0 (SPSS; Chicago, IL, USA) and Medcalc 19.1.7 (MedCals Software Ltd; Ostend, Belgium). *P*<0.05 was considered statistically significant.

## Result

### Patient demographic and clinical characteristics

Of 172 patients, 165 patients were included. As of 16 Mar 2020, 94 patients had clinical outcomes and 71 were follow-up lost without clinical outcome records due to hospital transfer (Figure 1). Of 94 patients, 81(86.2%) were discharged, 3(3.2%) were admitted to ICU, 4(4.3%) required mechanical ventilation, 6(6.4%) died. 31(38.3%) patients had complete absorption of lesions on CT after discharge. The median time from symptom onset to discharge was 21 (range, 10-41) days, and median times from symptom onset to being admitted to ICU, to requiring mechanical ventilation, and to death were 7 (range, 2-8) days, 8 (range, 8-49) days, and 33.5 (range, 7-39) days, respectively. The median times from symptom onset and from discharge to post-discharge CT scan were 37 (range, 14-58) days, 15 (range, 9-29) days, respectively.

**Figure 1.**
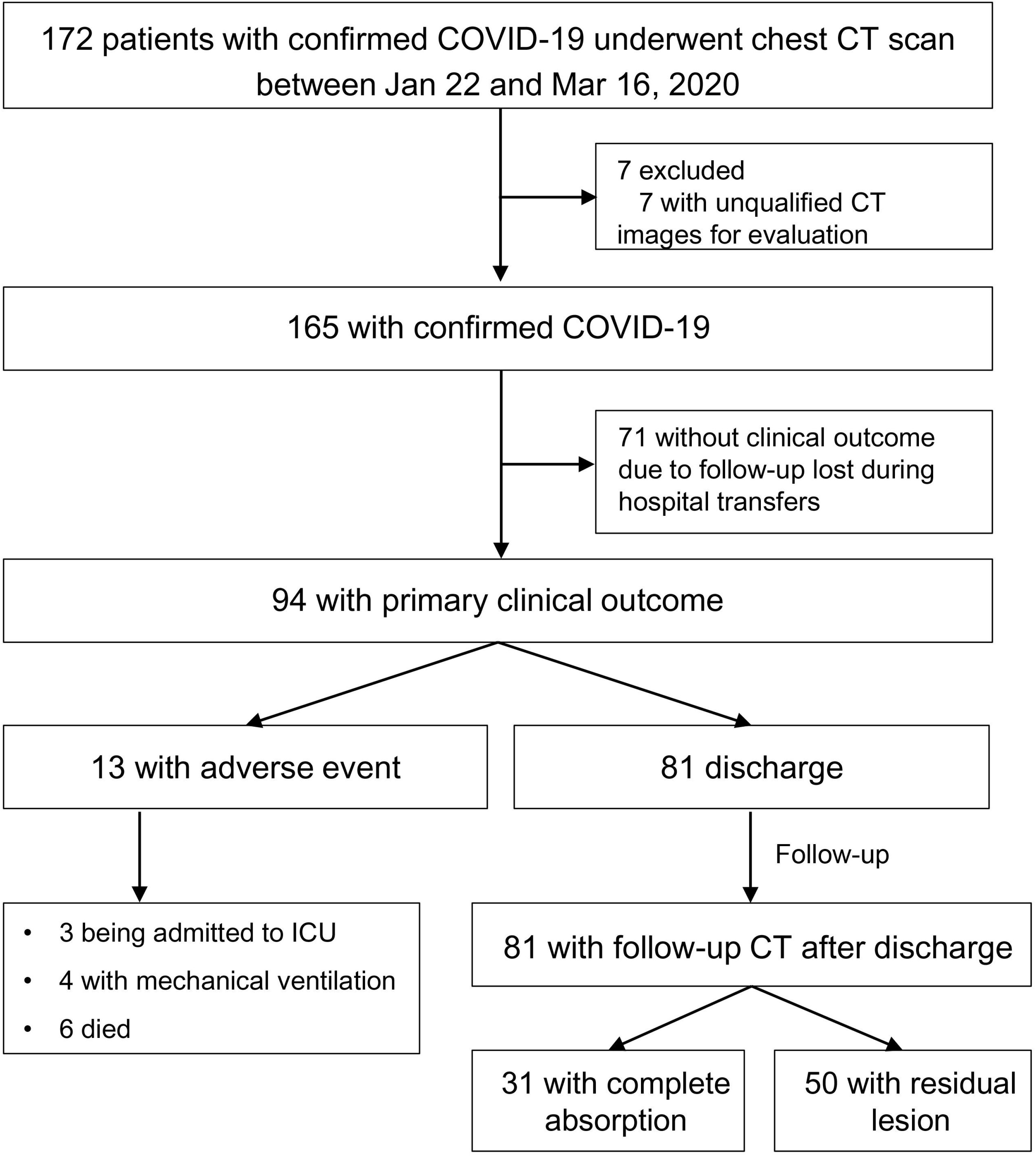
Study flow diagram. COVID-19 = coronavirus disease 2019, ICU = intensive care unit.

Patients were categorized into five CT patterns based on the baseline CT: 7(4.3%) were Pattern 0, 36(21.8%) were Pattern 1, 67(40.6%) were Pattern 2, 32(19.4%) were Pattern 3 and 23(13.9%) were Pattern 4. All the patients had 478 chest CT, 34(21.2%) had 1 CT, 41(23.6 %) had 2 CT, 39(23.7%) had 3 CT, and 51(31.5%) had more than 3 CT. The median time from symptom onset to baseline CT was 7 (range, 1-44) days.

Table 2 detailed the clinical characteristics and laboratory results of patients by CT pattern group. In the full cohort, the mean age was 49.5 (SD, 15.9; range, 4-89) years and there was no gender difference (91 [55.2%] men, 74[44.8%] women). Significant differences between pattern groups were found in age, sex distribution, disease severity, comorbidity, CT findings and laboratory results (all *P*<0.05). Significant differences were also observed in multiple comparisons between any two patterns in one or more than one terms of age, sex distribution, disease severity, comorbidity, CT findings and laboratory results (all *P*<0.017).

**Table 2.** Characteristics of COVID-19 pneumonia patients with various CT patterns.

| Characteristic | All<br>(n=165) | Pattern 0<br>(n=7) | Pattern 1<br>(n=36) | Pattern 2<br>(n=67) | Pattern 3<br>(n=32) | Pattern 4<br>(n=23) | P value | Pattern 0 | Pattern 1 | Pattern 2 | Pattern 3 |
| --- | --- | --- | --- | --- | --- | --- | --- | --- | --- | --- | --- |
|  |  |  |  |  |  |  |  | vs. | vs. | vs. | vs. |
|  |  |  |  |  |  |  |  | Pattern 1<br>P value | Pattern 2<br>P value | Pattern 3<br>P value | Pattern 4<br>P value |
| Age (years) <sup>a</sup> | 49.5±15.9 | 39.7±13.7 | 47.4±16.5 | 43.9±14.7 | 56.7±11.1 | 61.7±14.7 | <0.001 | 0.253 | 0.266 | <0.001<sup>†</sup> | 0.158 |
| Male Sex | 91(55.2) | 3(42.9) | 26(72.2) | 28(41.8) | 17(53.1) | 17(73.9) | 0.012 | 0.129 | 0.003<sup>†</sup> | 0.289 | 0.118 |
| Disease severity |  |  |  |  |  |  | <0.001 | 0.294 | 0.949 | <0.001<sup>†</sup> | 0.014 |
| Mild | 111(67.3) | 7(100) | 31(86.1) | 58(86.6) | 13(40.6) | 2(8.7) |  |  |  |  |  |
| Severe | 44(26.7) | 0 | 5(13.9) | 9(13.4) | 16(50.0) | 14(60.9) |  |  |  |  |  |
| Critical illness | 10(6.0) | 0 | 0 | 0 | 3(9.4) | 7(30.4) |  |  |  |  |  |
| Comorbidity | 101(61.2) | 2(28.6) | 8(22.2) | 21(31.3) | 18(56.2) | 15(65.2) | 0.002 | 0.716 | 0.326 | 0.018 | 0.503 |
| Clinical symptom on admission |  |  |  |  |  |  |  |  |  |  |  |
| Fever | 140(84.8) | 4(57.1) | 26(72.2) | 60(89.6) | 28(87.5) | 22(95.7) | 0.020 | 0.655 | 0.024 | 0.743 | 0.387 |
| Fatigue | 30(18.2) | 3(42.9) | 2(5.6) | 11(16.4) | 5(15.6) | 9(39.1) | 0.008 | 0.024 | 0.133 | 0.920 | 0.048 |
| Pharyngalgia | 18(10.9) | 2(28.6) | 4(11.1) | 9(13.4) | 2(6.3) | 1(4.3) | 0.347 | 0.248 | >0.999 | 0.495 | >0.999 |
| Headache | 6(3.6) | 0 | 2(5.6) | 4(6.0) | 0 | 0 | 0.602 | >0.999 | >0.999 | 0.301 | -- |
| Cough | 96(58.2) | 4(57.1) | 16(44.4) | 42(62.7) | 17(53.1) | 17(73.9) | 0.195 | 0.687 | 0.075 | 0.365 | 0.118 |
| Expectoration | 36(21.8) | 1(14.3) | 6(16.7) | 18(26.9) | 3(9.4) | 8(34.8) | 0.129 | >0.999 | 0.243 | 0.046 | 0.038 |
| Chest congestion/breath shortness | 34(20.6) | 0 | 2(5.6) | 9(13.4) | 13(40.6) | 10(43.5) | <0.001 | >0.999 | 0.321 | 0.002<sup>†</sup> | 0.832 |
| Muscle soreness | 8(4.8) | 0 | 2(5.6) | 4(6.0) | 1(3.1) | 1(4.3) | >0.999 | >0.999 | >0.999 | >0.999 | >0.999 |
| Nausea and vomiting | 1(0.6) | 0 | 1(2.8) | 0 | 0 | 0 | >0.999 | -- | >0.999 | >0.999 | -- |
| Diarrhea | 4(2.4) | 0 | 0 | 2(3.0) | 1(3.1) | 1(4.3) | 0.735 | -- | 0.541 | >0.999 | >0.999 |
| No symptom | 5(3.0) | 0 | 3(8.3) | 2(3.0) | 0 | 0 | 0.365 | >0.999 | 0.340 | >0.999 | -- |
| Laboratory test on admission |  |  |  |  |  |  |  |  |  |  |  |
| Lymphocyte percentage (%) |  |  |  |  |  |  | <0.001 | 0.280 | 0.097 | 0.004<sup>†</sup> | 0.836 |
| <20 | 62(38.0) | 0 | 6(16.7) | 21(31.8) | 20(62.5) | 15(65.2) |  |  |  |  |  |
| ≥20 | 101(62.0) | 6(100) | 30(83.3) | 45(68.2) | 12(37.5) | 8(34.8) |  |  |  |  |  |
| Monocyte percentage (%) |  |  |  |  |  |  | 0.315 | 0.414 | 0.085 | 0.102 | 0.261 |
| >10 | 39(24.5) | 1(16.7) | 12(33.3) | 12(18.2) | 10(33.3) | 4(19.0) |  |  |  |  |  |
| ≤10 | 120(75.5) | 5(83.3) | 24(66.7) | 54(81.8) | 20(66.7) | 17(81.0) |  |  |  |  |  |
| Leukocyte count (10<sup>9</sup>/L) |  |  |  |  |  |  | 0.062 | 0.167 | 0.570 | 0.924 | 0.014 |
| <3.5 | 40(24.5) | 0 | 9(25.0) | 20(30.3) | 10(31.2) | 1(4.3) |  |  |  |  |  |
| ≥3.5 | 123(75.5) | 6(100) | 27(75.0) | 46(69.7) | 22(68.8) | 22(95.7) |  |  |  |  |  |
| Alanine Aminotransferase (U/L) |  |  |  |  |  |  | 0.065 | 0.554 | 0.102 | 0.200 | 0.945 |
| >50 | 28(17.4) | 0 | 2(5.6) | 11(16.9) | 9(28.1) | 6(27.3) |  |  |  |  |  |
| ≤50 | 133(82.6) | 6(100) | 34(94.4) | 54(83.1) | 23(71.9) | 16(72.7) |  |  |  |  |  |
| Aspartate Aminotransferase (U/L) |  |  |  |  |  |  | 0.122 | 0.328 | 0.035 | 0.702 | 0.583 |
| >40 | 32(19.9) | 1(16.7) | 2(5.6) | 14(21.5) | 8(25.0) | 7(31.8) |  |  |  |  |  |
| ≤40 | 129(80.1) | 5(83.3) | 34(94.4) | 51(78.5) | 24(75.0) | 15(68.2) |  |  |  |  |  |
| Creatine kinase (U/L) |  |  |  |  |  |  | **0.014** | 0.014 | 0.022 | 0.429 | 0.038 |
| >310 | 18(11.8) | 1(16.7) | 0 | 9(13.6) | 2(7.7) | 6(31.5) |  |  |  |  |  |
| ≤310 | 134(88.2) | 5(83.3) | 35(100) | 57(86.4) | 24(92.3) | 13(68.4) |  |  |  |  |  |
| Neutrophil percentage (%) |  |  |  |  |  |  | **<0.001** | 0.391 | 0.080 | 0.232 | 0.043 |
| >75 | 48(29.4) | 0 | 4(11.1) | 17(25.8) | 12(37.5) | 15(65.2) |  |  |  |  |  |
| ≤75 | 115(70.6) | 6(100) | 32(88.9) | 49(74.2) | 20(62.5) | 8(34.8) |  |  |  |  |  |
| C-reactive protein (mg/L) |  |  |  |  |  |  | **0.002** | 0.130 | 0.245 | 0.356 | 0.055 |
| >10 | 96 (63.6) | 1(16.7) | 17(50.0) | 38(62.3) | 23(71.9) | 17(94.4) |  |  |  |  |  |
| ≤10 | 55(36.4) | 5(83.3) | 17(50.0) | 23(37.7) | 9(28.1) | 1(5.6) |  |  |  |  |  |
| Hemoglobin (g/L) |  |  |  |  |  |  | 0.494 | 0.873 | 0.976 | 0.684 | 0.251 |
| <130 | 35(22.4) | 1(16.7) | 7(19.4) | 13(19.7) | 7(23.3) | 7(38.9) |  |  |  |  |  |
| ≥130 | 121(77.6) | 5(83.3) | 29(80.6) | 53(80.3) | 23(76.7) | 11(61.1) |  |  |  |  |  |
| CT findings on admission |  |  |  |  |  |  |  |  |  |  |  |
| CT signs |  |  |  |  |  |  |  |  |  |  |  |
| GGO only | 28(17.0) | 0 | 13(36.1) | 12(17.9) | 2(6.3) | 1(4.3) | **0.005** | -- | 0.040 | 0.215 | >0.999 |
| Consolidation | 17(10.3) | 0 | 5(13.9) | 6(9.0) | 3(9.4) | 3(13.0) | 0.880 | -- | 0.510 | >0.999 | 0.686 |
| GGO and consolidation | 51(30.9) | 0 | 10(27.8) | 16(23.9) | 10(31.3) | 15(65.2) | **0.002** | -- | 0.664 | 0.436 | **0.013** |
| Linear opacity | 0 | 0 | 0 | 0 | 0 | 0 | -- | -- | -- | -- | -- |
| GGO and linear opacity | 7(4.2) | 0 | 2(5.6) | 3(4.5) | 2(6.3) | 0 | 0.839 | -- | >0.999 | 0.657 | 0.504 |
| Consolidation and linear opacity | 5(3.0) | 0 | 1(2.8) | 4(6.0) | 0 | 0 | 0.618 | -- | 0.665 | 0.301 | -- |
| Three mixed signs | 50(30.3) | 0 | 5(13.9) | 26(38.8) | 15(46.9) | 4(17.4) | **0.003** | -- | **0.009<sup>†</sup>** | 0.446 | 0.023 |
| Lobe involvement |  |  |  |  |  |  | **<0.001** | -- | 0.121 | **0.008<sup>†</sup>** | 0.632 |
| Number of lobe affected<3 | 52(31.5) | 7(100) | 18(50.0) | 23(34.3) | 3(9.4) | 1(4.3) |  |  |  |  |  |
| Number of lobe affected≥3 | 113(68.5) | 0 | 18(50.0) | 44(65.7) | 29(90.6) | 22(95.7) |  |  |  |  |  |
| CT severity score <sup>a</sup> | 6.0±4.4 | 0 | 3.3±2.1 | 4.7±2.7 | 7.5±2.8 | 14.0±2.9 | **<0.001** | **<0.001<sup>†</sup>** | **0.005<sup>†</sup>** | **<0.001<sup>†</sup>** | **<0.001<sup>†</sup>** |

Note: Unless otherwise indicated, data are reported as the number of patients, with percentages in parentheses. a, data are reported as the mean±standard derivation. b, more than 91.5% of patients had all laboratory tests and a few were lack of one or two indicators. †, Significance at $P<0.0125$ with Bonferroni correction. Abbreviations: GGO = ground glass opacity; Three mixed signs = GGO, consolidation and linear opacity.

### Evolution of COVID-19 pneumonic CT pattern with disease progression

Chi-square tests for trend indicated that as disease progresses from 1 to >3 weeks, proportions of Pattern 1 and 2 remarkably decreased, while those of Pattern 3 and 4 increased (all *P*<0.01). With regard to evolution of CT pattern, Pattern 0 to 2 showed a remarkable evolution with overlaps of progression and downgrade within 3 weeks after symptom onset, and mostly remained the same thereafter. Pattern 3 and 4 showed a remarkable evolution (progression or downgrade) within 2 weeks, and most of them remained afterwards. (Figure 2)

**Figure 2.**
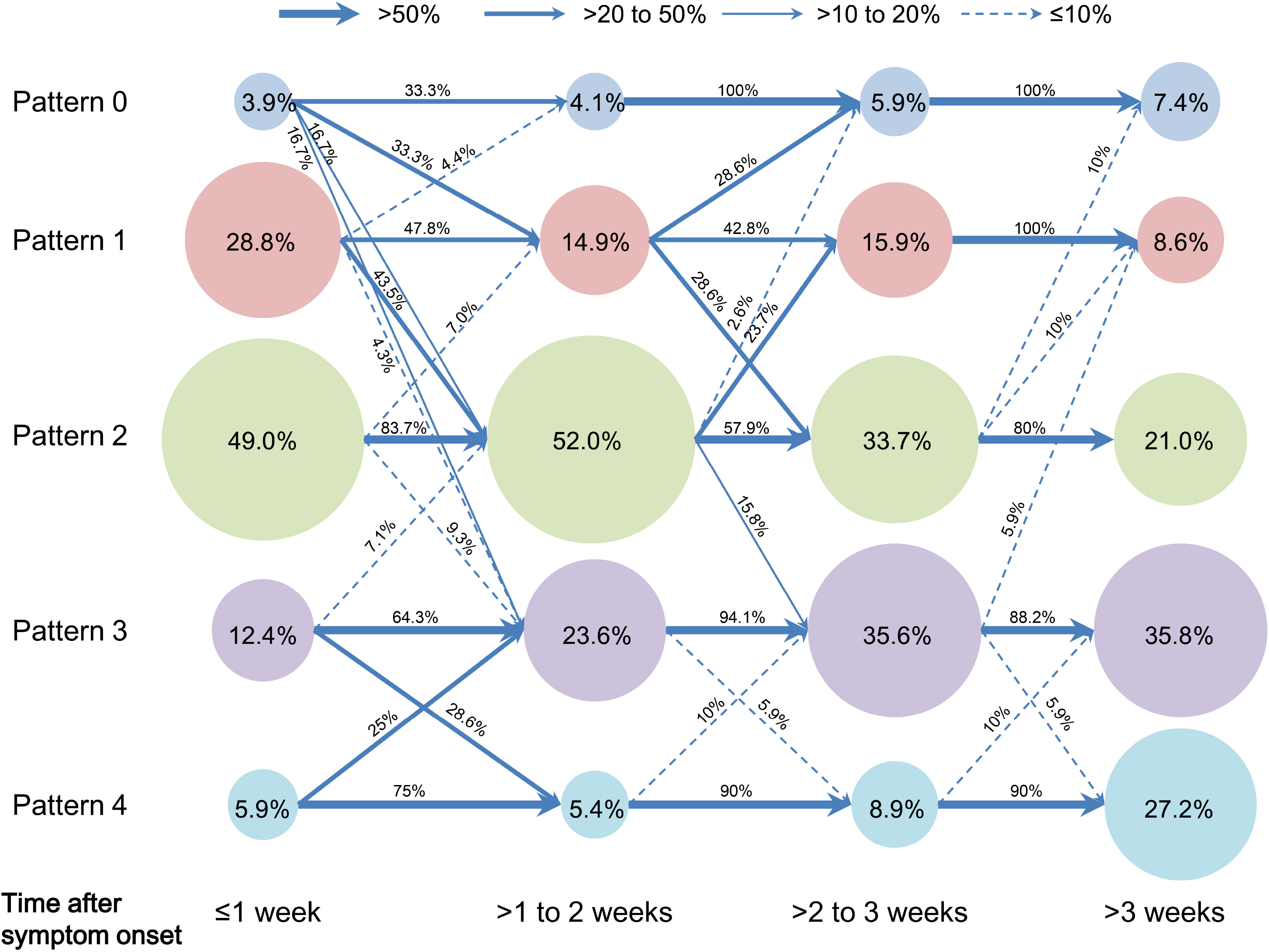
Evolution of proportions of COVID-19 pneumonic CT pattern with the disease progression. Pattern data were designated to four time groups according to the time from symptom onset to CT scan: ≤1 week (CT pattern number=153), >1 to 2 weeks (CT pattern number =147), >2 to 3 weeks (CT pattern number =101) and >3 weeks (CT pattern number =77). The circular area indicated the proportion of CT pattern in each time group, e.g. proportions of Pattern 0 to 4 were 3.9%, 28.8%, 49.0%, 12.4% and 5.9% during 1 week after symptom onset, respectively. Arrow line indicated the evolution of each CT pattern from a time group to the following, e.g. 33.3% of Pattern 1 progressed to Pattern 2 from 1 to 2 weeks after symptom onset; here four arrow line style denoted the categorization of evolution proportion, i.e. >50% (thick solid line), >20 to 50% (medium solid line), >10 to 20% (thin solid line) and ≤10% (dashed line).

Figure 3-6 presented CT findings with disease progression in Pattern 1 to 4 cases. Pattern 1 and 2 showed limited progression with increasing density and size of lesions from 1 to 2 weeks after onset, while had complete absorption subsequently. Pattern 3 showed a fast progression from patchy GGO to extensively mixed GGO and consolidation within 2 weeks, and subsequently turned into mixed GGO and linear opacities. Pattern 4 showed a considerably fast progression to diffusely mixed consolidation and interlobular septal thickening in both lungs and had adverse outcome within 1 week.

**Figure 3.**
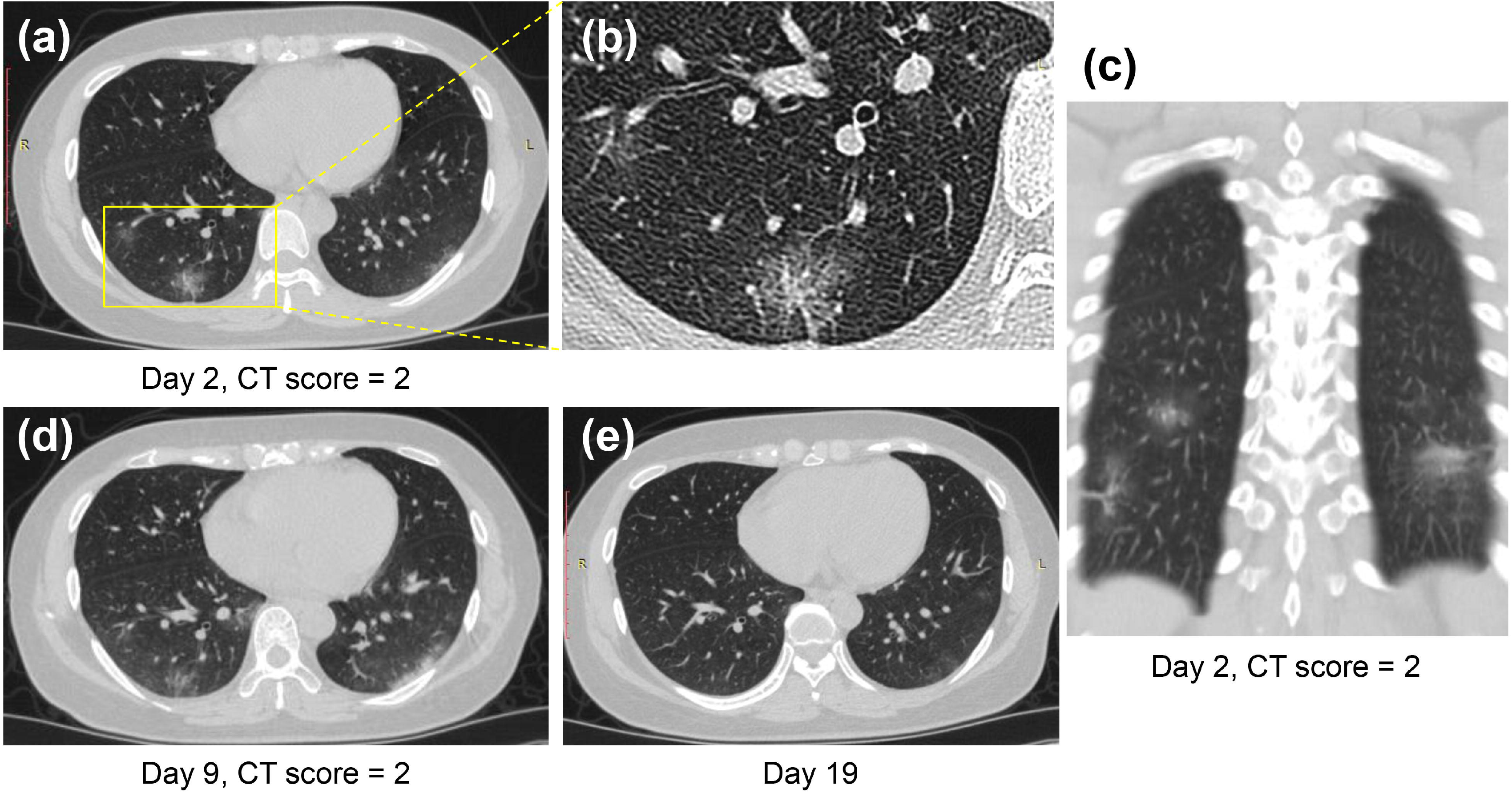
CT Pattern 1 (bronchopneumonia pattern) in a 38-year-old woman with COVID-19 pneumonia who was admitted to hospital at day 2 after symptom onset. (A, B, C) Axial and coronal CT images demonstrate multifocal peribronchial ground-glass opacity (GGO) at day 2; Axial CT images demonstrate increasing density and size of lesions at day 9 (D) and subsequently complete absorption at day 19 (E).

**Figure 4.**
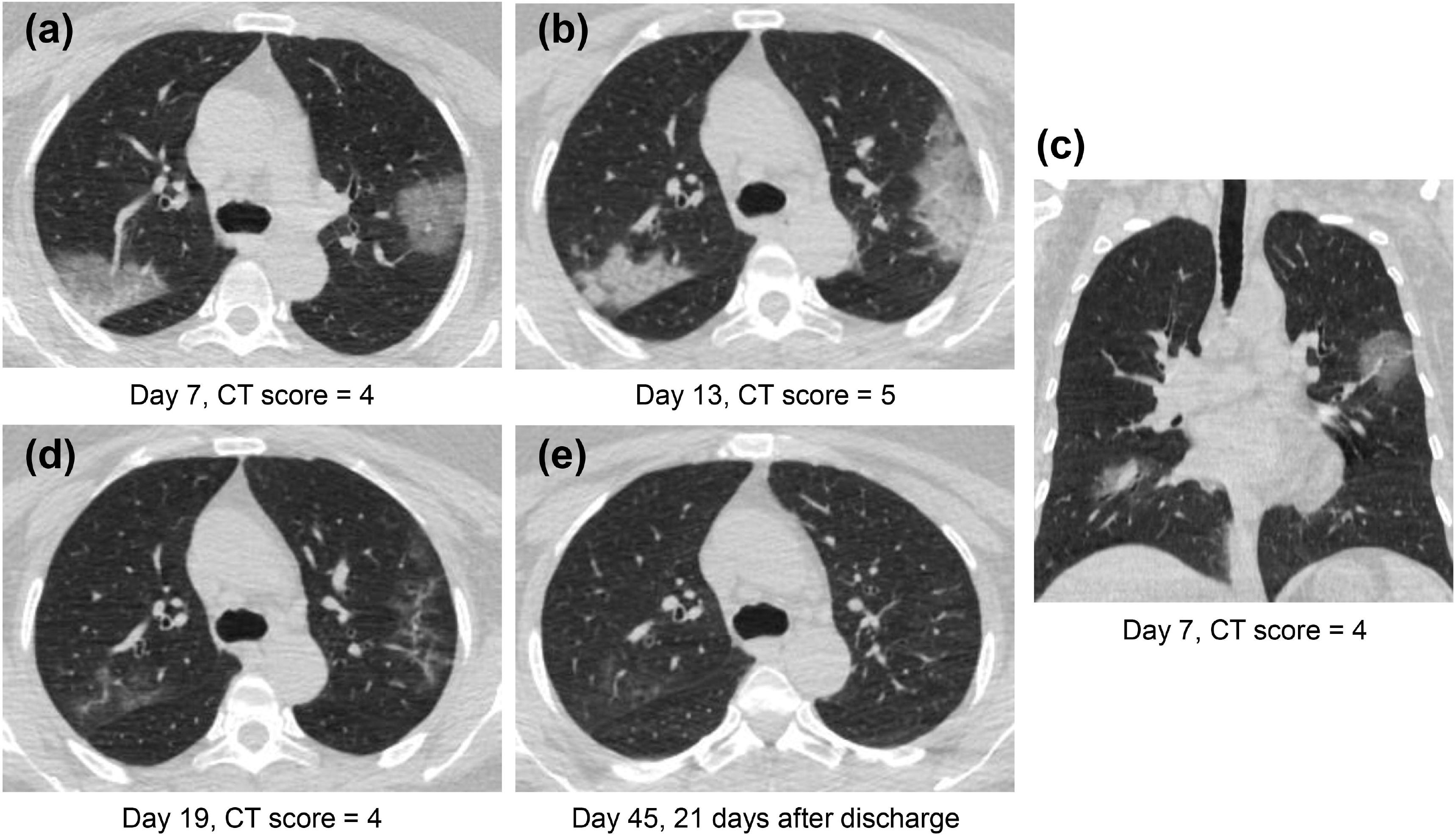
CT Pattern 2 (organizing pneumonia pattern) in a 49-year-old woman with COVID-19 pneumonia who was admitted to hospital at day 7 after symptom onset and discharged at day 24. (A, C) Axial and coronal CT images demonstrate multifocal ground-glass opacity (GGO), mixed GGO and consolidation at day 7; Axial CT images demonstrate consolidation at day 13 (B), subsequent absorption with mixed GGO and linear opacities at day 19 (D), and complete absorption at day 45 (E).

**Figure 5.**
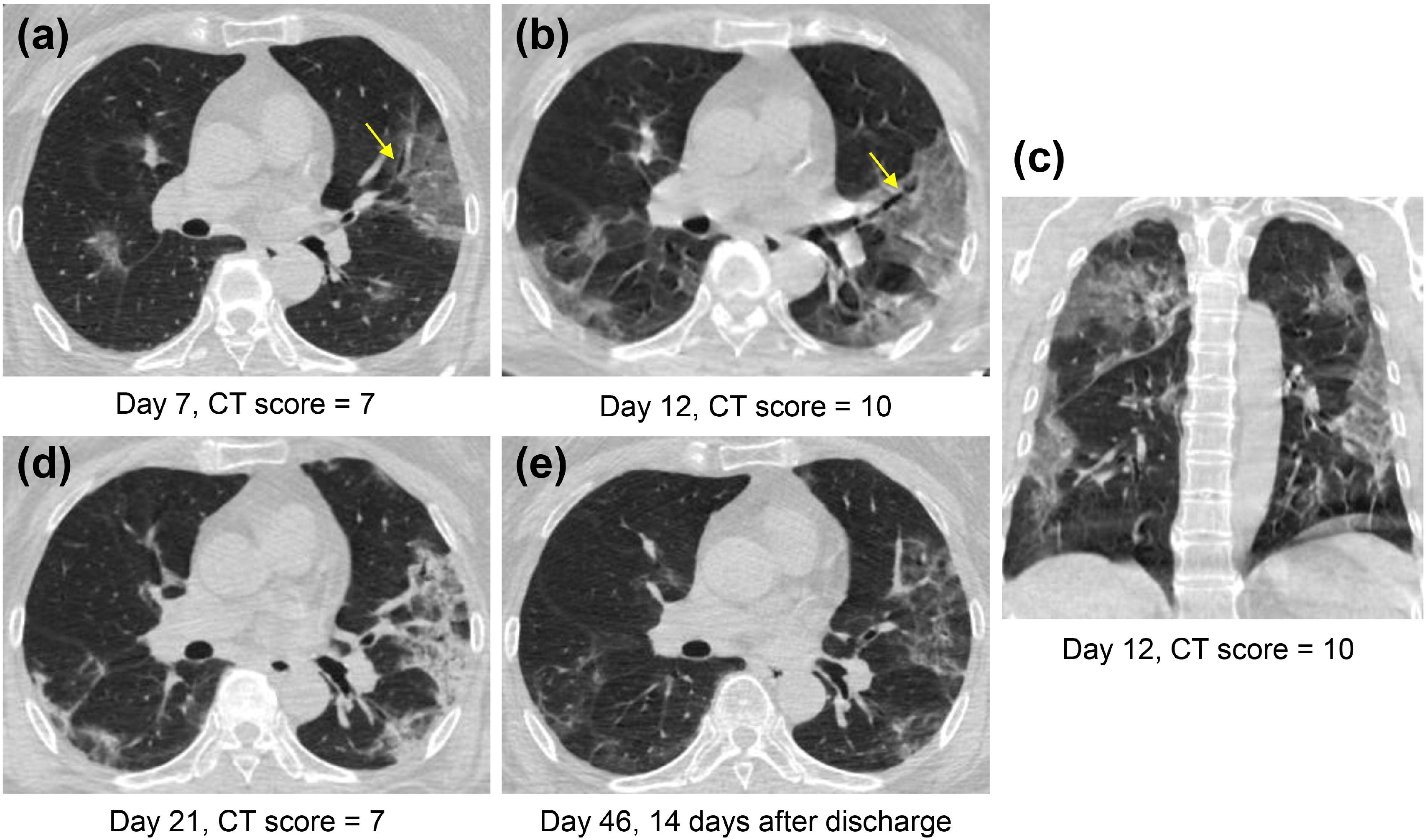
CT Pattern 3 (progressive organizing pneumonia pattern) in a 65-year-old woman with COVID-19 pneumonia who was admitted to hospital at day 7 after symptom onset and discharged at day 24. Axial and coronal CT images demonstrate a fast progression from patchy ground-glass opacity (GGO) with slight bronchial dilatation (arrow) at day 7 (A), to extensive GGO and consolidation with progressive bronchial dilatation (arrow) at day 12 (B, C); Axial CT images reveal that extensive GGO and consolidation turned into consolidation and reticulation at day 21 (D) and into mixed GGO and linear opacities at day 46 (E).

**Figure 6.**
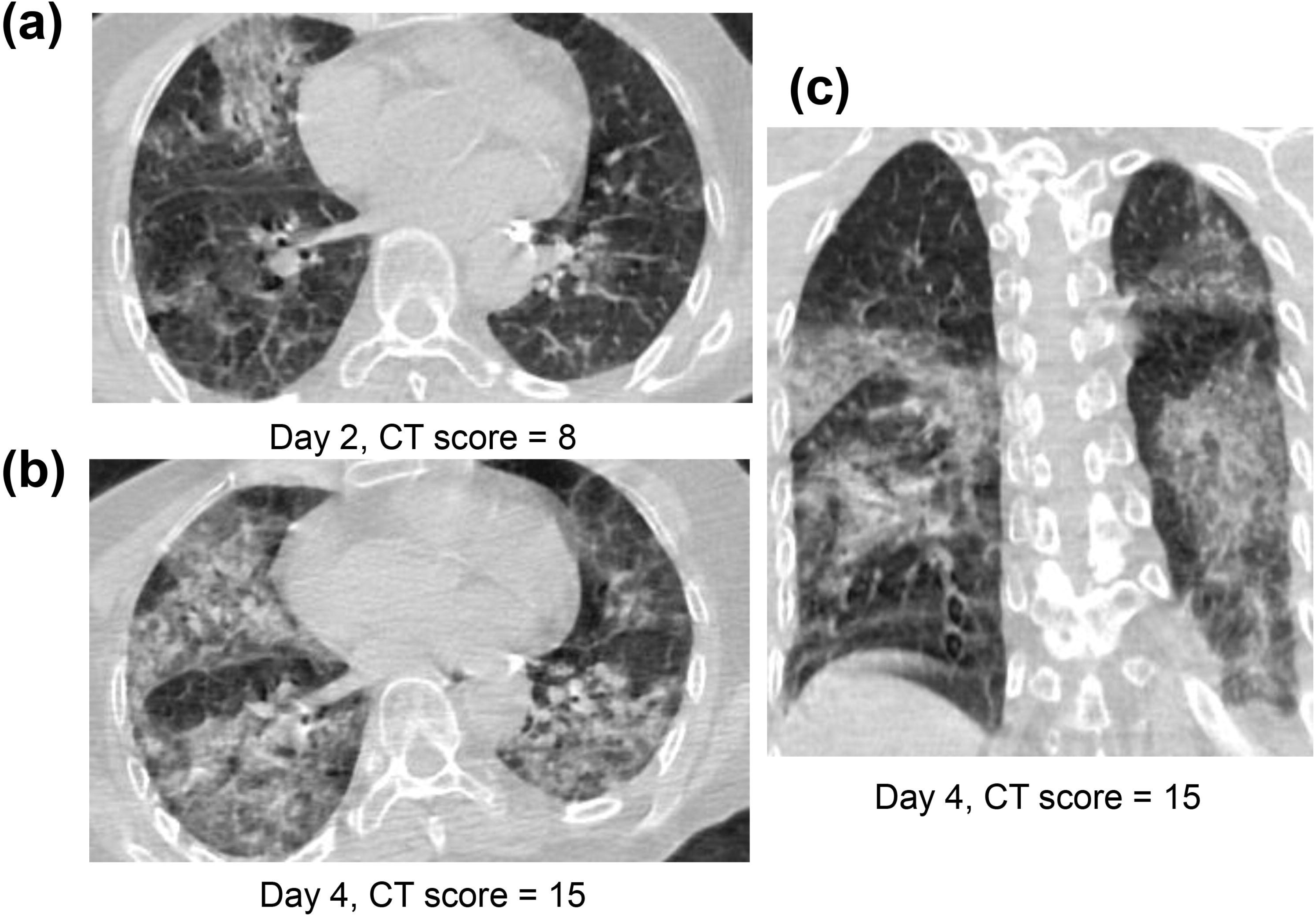
CT Pattern 4 (diffuse alveolar damage pattern) in an 82-year-old woman COVID-19 pneumonia and with history of cardiovascular disease and chronic obstructive pulmonary disease, who was admitted to intensive care unit with mechanical ventilation at day 7 after symptom onset. Axial CT images demonstrate a fast progression from mixed ground-glass opacity (GGO) and consolidation at day 2 (A) to a geographic distribution of mixed consolidation and interlobular septal thickening at day 4 (B); (C) Coronal CT image demonstrates mixed consolidation and interlobular septal thickening with diffused distribution of both lungs.

### Prognostic significance of pneumonic CT pattern in COVID-19

Table E1 detailed the clinical, laboratory and CT imaging characteristics of patients in clinical outcome and pulmonary sequelae on CT. Significant differences between discharge and adverse outcome were found in age, disease severity, comorbidity, laboratory results, CT pattern and CT score (all *P*<0.05). For pulmonary sequelae, significant differences between complete absorption and residuals were found in age, elevated neutrophil percentage, elevated C-reactive protein, CT pattern and CT score (all *P*<0.05).

### Correlations of CT pattern with clinical outcomes

Univariate Cox proportional-hazards regression indicated that CT Pattern 4 (Hazard ratio 36.67, 95%CI 4.38-307.25, *P*=0.001) significantly correlated with adverse outcomes. Besides, age ≥65 years (9.39, 2.38-37.11, *P*=0.001), comorbidity (4.14, 1.09-15.71, *P*=0.037), severe or critical illness (4.62, 2.04-10.46, *P*<0.001), presence of fatigue (3.62, 1.16-11.28, *P*=0.027) and chest congestion and/or shortness of breath (3.81, 1.19-12.18, *P*=0.024), neutrophil percentage >75% (14.12, 1.75-114.21, *P*=0.013), CT score ≥10 (11.66, 2.31-58.75, *P*=0.003) were associated with adverse outcomes (Table 3). Multivariate analysis indicated that after controlling for age, Pattern 4 was found to be an independent risk factor for adverse outcomes (18.90, 1.91-186.60, *P*=0.012) (Figure 7).

**Table 3.** Risk factors associated with adverse outcome in patients with COVID-19 pneumonia.

| Variable | Stratification | Univariate analysis |  |  | Multivariate analysis |  |  |
| --- | --- | --- | --- | --- | --- | --- | --- |
|  |  | HR | 95%CI | P value | HR | 95%CI | P value |
| Age (years) | ≥65 vs. <65 (Ref.) | 9.39 | 2.38-37.11 | **0.001** | 3.04 | 0.74-12.56 | 0.124 |
| Sex | Male vs. female (Ref.) | 0.86 | 0.27-2.77 | 0.805 |  |  |  |
| Comorbidity | Yes vs. No (Ref.) | 4.14 | 1.09-15.71 | **0.037** |  |  |  |
| Disease severity | Severe, critical illness vs. Mild (Ref.) | 4.62 | 2.04-10.46 | **<0.001** |  |  |  |
| Laboratory test at admission |  |  |  |  |  |  |  |
| Lymphocyte percentage (%) | <20 vs. ≥20 (Ref.) | 1.00 | 0.24-4.16 | 0.998 |  |  |  |
| Monocyte percentage (%) | >10 vs. ≤10 (Ref.) | 0.33 | 0.04-2.60 | 0.294 |  |  |  |
| Leukocyte count (10<sup>9</sup>/L) | <3.5 vs. ≥3.5 (Ref.) | 0.03 | 0-76.60 | 0.390 |  |  |  |
| Alanine Aminotransferase (U/L) | >50 vs. ≤50 (Ref.) | 0.82 | 0.21-3.16 | 0.820 |  |  |  |
| Aspartate Aminotransferase (U/L) | >40 vs. ≤40 (Ref.) | 2.01 | 0.63-6.40 | 0.239 |  |  |  |
| Creatine kinase (U/L) | >310 vs. ≤310 (Ref.) | 3.39 | 0.87-13.18 | 0.078 |  |  |  |
| Neutrophil percentage (%) | >75 vs. ≤75 (Ref.) | 14.12 | 1.75-114.21 | **0.013** |  |  |  |
| C-reactive protein (mg/L) | >10 vs. ≤10 (Ref.) | 53.87 | 0.12-2.5×10<sup>4</sup> | 0.203 |  |  |  |
| Hemoglobin (g/L) | <130 vs. ≥130 (Ref.) | 0.69 | 0.17-2.83 | 0.606 |  |  |  |
| CT findings |  |  |  |  |  |  |  |
| GGO only | Yes vs. No (Ref.) | 2.79 | 0.34-23.19 | 0.343 |  |  |  |
| Consolidation | Yes vs. No (Ref.) | 0.04 | 0-6781 | 0.607 |  |  |  |
| GGO and consolidation | Yes vs. No (Ref.) | 3.24 | 0.93-11.27 | 0.065 |  |  |  |
| Linear opacity | Yes vs. No (Ref.) | -- | -- | -- |  |  |  |
| GGO and linear opacity | Yes vs. No (Ref.) | 0.04 | 0-2.3×10<sup>4</sup> | 0.641 |  |  |  |
| Consolidation and linear opacity | Yes vs. No (Ref.) | 0.05 | 0-1.7×10<sup>6</sup> | 0.730 |  |  |  |
| Three mixed signs | Yes vs. No (Ref.) | 0.47 | 0.13-1.74 | 0.255 |  |  |  |
| Number of lobe affected | >3 vs. ≤3 (Ref.) | 4.86 | 0.59-39.77 | 0.141 |  |  |  |
| CT severity score | ≥10 vs. <10 (Ref.) | 11.66 | 2.31-58.75 | **0.003** |  |  |  |
| CT pattern | Pattern 4 vs. Pattern 0-3 (Ref.) | 36.67 | 4.38-307.25 | **0.001** | 18.90 | 1.91-186.60 | **0.012** |

Note: Ref. refers to the stratification of variable as reference in the Cox hazard-proportional regression analysis.
Abbreviations: HR = hazard ratio; 95% CI = 95% confidence interval; GGO = ground glass opacity; Three mixed signs = GGO, consolidation and linear opacity.

**Figure 7.**
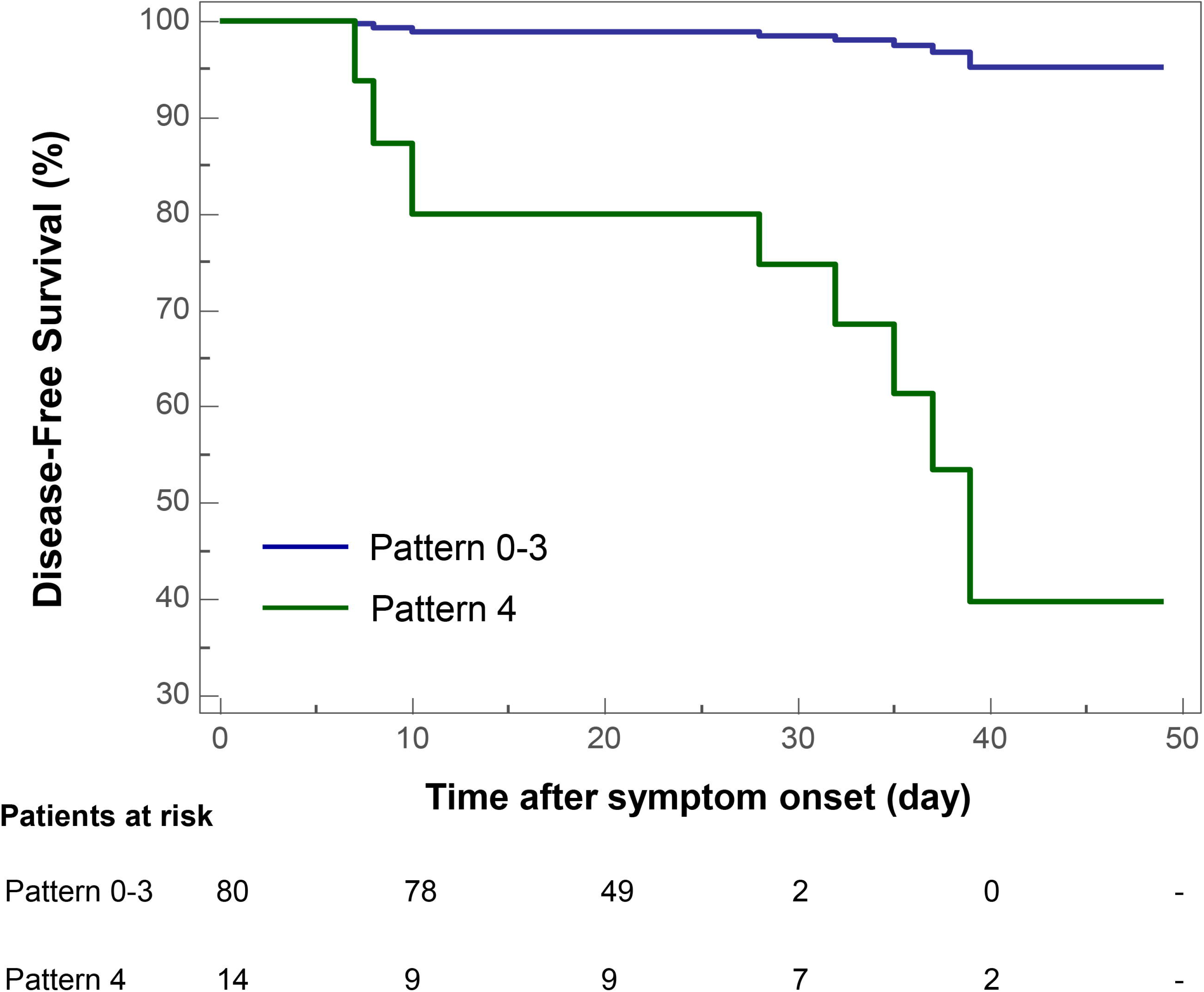
Kaplan-Meier curve plots showing time from symptom onset to adverse outcome events (admission to intensive care unit, use of mechanical ventilation, or death) by categories of COVID-19 pneumonic CT pattern (Pattern 4 vs. Pattern 0-3 as reference).

### Correlations of CT pattern with pulmonary sequelae on CT after discharge

By univariate Cox proportional-hazards regression, it was found that CT Pattern 3 or 4 (0.23, 0.07-0.78, *P*=0.017) were significantly related with pulmonary sequelae. Beyond, significant factors included age ≥45 years (0.36, 0.15-0.88, *P*=0.025), C-reactive protein concentration >10 mg/L (0.28, 0.12-0.65, *P*=0.003), number of lobe affected >3 (0.34, 0.16-0.71, *P*=0.005), CT score ≥4 (0.32, 0.15-0.65, *P*=0.002) (Table 4). The multivariate analysis showed that Pattern 3 or 4 (0.26, 0.08-0.88, *P*=0.030) and C-reactive protein (0.31, 0.13-0.72, *P*0.006) were two independent factors associated with pulmonary residuals (Figure 8).

**Table 4.** Risk factors associated with pulmonary sequelae of lesion resolution at 2-3 weeks after discharge in patients with COVID-19 pneumonia.

| Variable | Stratification | Univariate analysis |  |  | Multivariate analysis |  |  |
| --- | --- | --- | --- | --- | --- | --- | --- |
|  |  | HR | 95%CI | P value | HR | 95%CI | P value |
| Age (yr) | ≥45 vs. <45 (Ref.) | 0.36 | 0.15-0.88 | **0.025** |  |  |  |
| Sex | Male vs. Female (Ref.) | 1.09 | 0.53-2.25 | 0.806 |  |  |  |
| Comorbidity | Yes vs. No (Ref.) | 0.46 | 0.18-1.21 | 0.116 |  |  |  |
| Disease severity | Severe vs. Mild (Ref.) | 0.87 | 0.12-6.43 | 0.893 |  |  |  |
| Laboratory test at admission |  |  |  |  |  |  |  |
| Lymphocyte percentage (%) | <20 vs. ≥20 (Ref.) | 0.50 | 0.22-1.13 | 0.094 |  |  |  |
| Monocyte percentage (%) | >10 vs. ≤10 (Ref.) | 1.94 | 0.92-4.09 | 0.082 |  |  |  |
| Leukocyte count (10<sup>9</sup>/L) | <3.5 vs. ≥3.5 (Ref.) | 0.96 | 0.39-2.38 | 0.928 |  |  |  |
| Alanine Aminotransferase (U/L) | >50 vs. ≤50 (Ref.) | 0.50 | 0.17-1.46 | 0.202 |  |  |  |
| Aspartate Aminotransferase (U/L) | >40 vs. ≤40 (Ref.) | 0.69 | 0.27-1.81 | 0.451 |  |  |  |
| Creatine kinase (U/L) | >310 vs. ≤310 (Ref.) | 0.50 | 0.12-2.12 | 0.349 |  |  |  |
| Neutrophil percentage (%) | >75 vs. ≤75 (Ref.) | 0.32 | 0.10-1.06 | 0.062 |  |  |  |
| C-reactive protein (mg/L) | >10 vs. ≤10 (Ref.) | 0.28 | 0.12-0.65 | **0.003** | 0.31 | 0.13-0.72 | **0.006** |
| Hemoglobin (g/L) | <130 vs. ≥130 (Ref.) | 0.36 | 0.09-1.54 | 0.169 |  |  |  |
| CT findings |  |  |  |  |  |  |  |
| GGO only | Yes vs. No (Ref.) | 1.14 | 0.34-3.84 | 0.827 |  |  |  |
| Consolidation | Yes vs. No (Ref.) | 2.89 | 1.08-7.72 | **0.035** |  |  |  |
| GGO and consolidation | Yes vs. No (Ref.) | 1.02 | 0.45-2.28 | 0.969 |  |  |  |
| Linear opacity | Yes vs. No (Ref.) | -- | -- | -- |  |  |  |
| GGO and linear opacity | Yes vs. No (Ref.) | 0.88 | 0.21-3.71 | 0.856 |  |  |  |
| Consolidation and linear opacity | Yes vs. No (Ref.) | 0.89 | 0.12-6.59 | 0.911 |  |  |  |
| Three mixed signs | Yes vs. No (Ref.) | 0.52 | 0.24-1.13 | 0.098 |  |  |  |
| Number of lobe affected | >3 vs. ≤3 (Ref.) | 0.34 | 0.16-0.71 | **0.005** |  |  |  |
| CT severity score | ≥4 vs. <4 (Ref.) | 0.32 | 0.15-0.65 | **0.002** |  |  |  |
| CT Pattern | Pattern 3,4 vs. Pattern 0-2 (Ref.) | 0.23 | 0.07-0.77 | **0.017** | 0.26 | 0.08-0.88 | **0.030** |

Note: Ref. refers to the stratification of variable as reference in the Cox hazard-proportional regression analysis.
Abbreviations: HR = hazard ratio; 95% CI = 95% confidence interval; GGO = ground glass opacity; Three mixed signs = GGO, consolidation and linear opacity.

**Figure 8.**
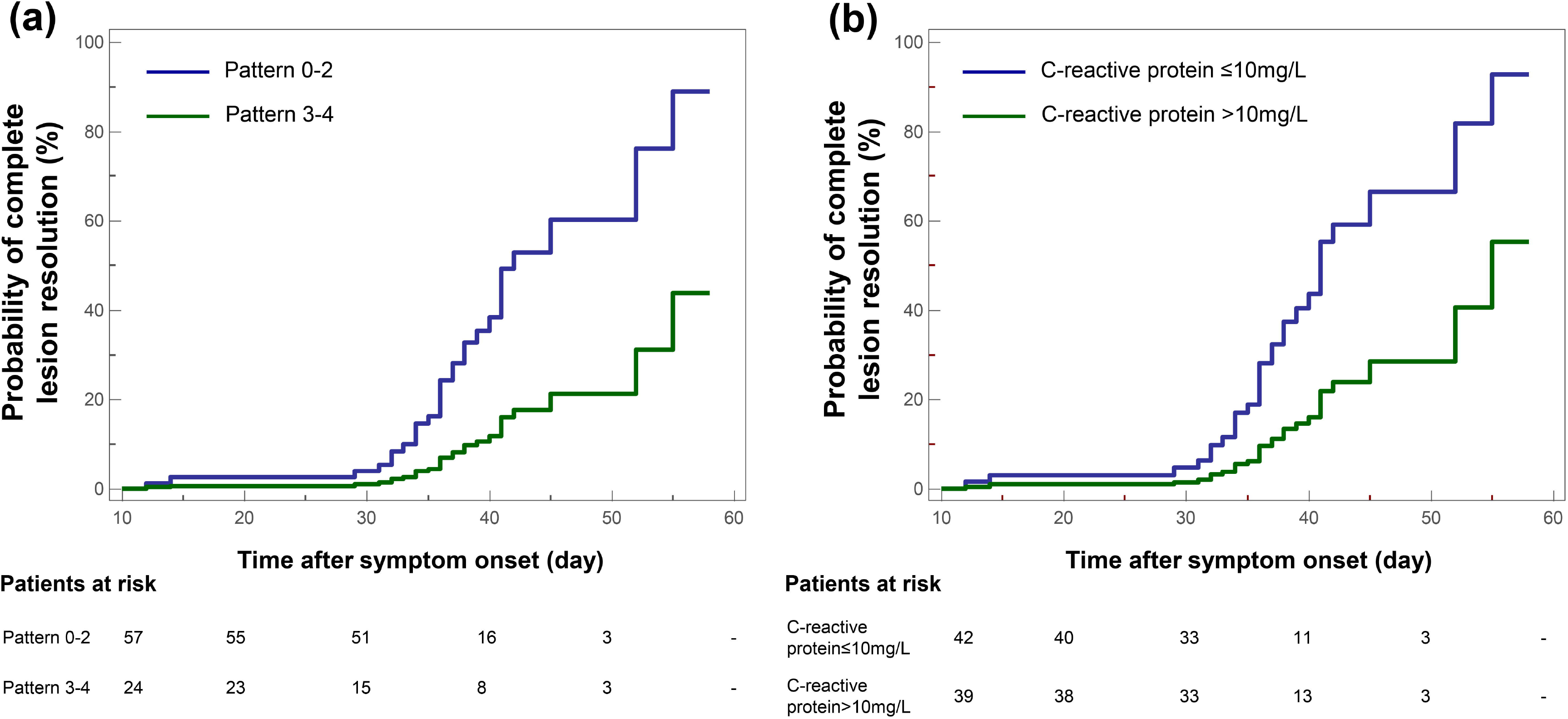
Kaplan-Meier curve plots showing time from symptom onset to complete resolution of pulmonary lesions by (A) categories of COVID-19 pneumonic CT pattern (Pattern 3-4 vs. Pattern 0-2 as reference), and (B) conditions of C-reactive protein.

## Discussion

By delineating the COVID-19 pneumonic CT patterns and their evolutional characteristics, this study aimed to determine their value in predicting adverse outcomes. Results indicated that CT Pattern 4 was associated with a higher rate of an adverse outcome after controlling for age; meanwhile, Pattern 3 and 4 showed remarkably more prevalence of pulmonary residuals on CT. Individual CT pattern for prognostic implication can be determined within 2 weeks after symptom onset due to the remarkable evolution of patterns before 2 weeks and subsequent stabilization or evolution without prognostic impacts.

A categorization of CT patterns was performed by detailing the extent of lung injury in COVID-19. Differently from prior categorization of bronchopneumonia, OP and DAD [17], lung involvement characterized by CT score was evaluated besides the CT signs. Specifically, receiver operating characteristic curve analysis estimated the cutoff CT scores of 6 and 10 in discriminations of Pattern 2 vs. 3 and Pattern 3 vs. 4, respectively (See more in Supplement). Pattern 1 was found to be linked with a good prognosis as well as Pattern 0. This resembled the prior reports of H1N1 pneumonia [17]. In contrast to other patterns, Pattern 1 may represent the early stage of COVID-19 pneumonia and can be considered as mild illness group. Pathologically, organization has been recognized as a common response in lung injury [15,23]. In this study, OP patterns accounted for 60% and the overall degree of lung injury in most cases especially for Pattern 2 was mild where reparative process and resolution of lesions seem to follow. Note that more prevalence of residuals may indicate a protracted disease course in Pattern 3. This may be related to the facts that Pattern 3 cases tend to be older with higher prevalence of comorbidity and decreased lymphocyte percentage. For Pattern 4, 85.7% cases had an adverse outcome. Pathologically, intraalveolar edema, fibrin, and variable cellular infiltrates with a hyaline membrane were observed in DAD [16,24]. Pattern 4 cases had more severe disease, more prevalence of elevated creatine kinase, neutrophil percentage and C-reactive protein than other patterns. It may be such facts that led to the higher rate of adverse outcomes in Pattern 4. Previous studies have demonstrated the residual fibrosis in 38% and 85% of DAD survivals, which may be related to barotrauma due to mechanical ventilation or oxygen toxicity [25]. Differently, fibrosis was not pathologically observed in COVID-19 death perhaps due to the short disease course of 15 days from onset to death [26]. A long-term follow up of discharged DAD patients who survived after mechanical ventilation or continuous high-flow oxygen therapy would be required to further understand the sequelae. All the above demonstrated the potentials of CT pattern categorization in predictive of prognosis in COVID-19.

Diverse evolutions with overlaps of progression and downgrading were found in Pattern 0-2 within 3 weeks and Pattern 3-4 within 2 weeks after onset. Most of them remained thereafter. It is noting that 28.6% of Pattern 1 progressed to Pattern 2 from 2 to 3 weeks. This evolution was consistent with prior report of acute and progressive characteristics of COVID-19 [11]. In addition, this progression from Pattern 1 to 2 after 2 weeks may reflect the organization regarding lung repair and would have good prognosis [15]. From the above, individual CT pattern for prognostic implication can be determined within 2 weeks after onset due to the remarkable evolution of patterns before 2 weeks and subsequent stabilization or evolution without prognostic impacts.

Univariate analysis indicated that age ≥65 years, presence of comorbidity (70% hypertension and diabetes mellitus), severe or critical illness, neutrophil percentage >75%, CT score ≥10, CT Pattern 4 were significantly related with adverse outcome. These findings echo the latest reports [7,8]. In details, a poor clinical outcome was associated with increased age (>65 years) and presence of comorbidity in COVID-19 patients [7,8]. Laboratory parameters including elevated levels of hypersensitive troponin I, leukocyte and neutrophil were also found to be predictive of a poor outcome [7-9]. By multivariate analysis, only Pattern 4 was associated with an adverse outcome after controlling age. In our cohort, most of Pattern 4 cases were age ≥65 years (64.3%), presence of comorbidity (71.4%) and critical illness (57.1%). This may be the underlying reason regarding Pattern 4 as only significant factor in multivariate analysis. This further enhanced the potential role of CT pattern in predicting the risks of adverse outcomes in COVID-19.

As for pulmonary sequelae, CT Pattern 3 or 4 and elevated C-reactive protein were two independent factors associated with pulmonary residuals on CT. Pattern 3 and 4 showed more prevalence of pulmonary residuals than others. This may be linked with more severe CT findings of these cases with more number of lobe affected and CT scores. In concert with MERS studies that radiological sequelae can remain at least 1 year after infection [27], our study found similar but slighter residuals mainly presenting with linear opacities and/or a few consolidation and GGO. Beyond, elevated C-reactive protein may indicate the state of tissue injury and/or acute inflammation. Continuous high levels of C-reactive protein in respiratory infections were suggestive of a risk indication of progression to a critical disease state [28]. In this regard, elevated C-reactive protein may be predictive of radiological sequelae. Prior studies indicated that radiological sequelae from SARS and MERS may suggest the abnormal or repaired lung function [27,29]. Despite the slight residuals in COVID-19, a long-term follow-up is required to further trace the resolution and associations with lung function.

This study had some limitations. The first was the small sample, especially for those with adverse outcomes. A larger sample is required to further verify the findings regarding the risk factors affecting the adverse outcome. Second, because discharged patients remained during the recovery and pulmonary CT residuals were unknown at the time of our analysis, a long-term follow-up is required to further trace the outcome of lesion absorption, as well as changes in lung functions. Last, multicenter data collection may lead to selective bias of patients with various CT patterns. Although no significance in univariate analysis (see more in Supplement), potential impacts from varying hospital, epicenter vs. non-epicenter should be considered in further studies.

In conclusion, CT pattern categorization of COVID-19 pneumonia based on chest CT within 2 weeks after symptom onset has prognostic significance. CT pattern 4 cases present high risk of admission to ICU, need for mechanical ventilation or death, while Pattern 3 and 4 signal likelihood of pulmonary residuals on CT. These findings would help early prognostic stratification of COVID-19 and facilitate the decision making for treatment strategy and optimal use of healthcare resources.

## Data Availability

The data that support the findings of this study are available on request from the corresponding author.

## Declarations

### Ethics approval

This study was approved by the Institutional Review Board of the First Affiliated Hospital of Xi’an Jiaotong University.

### Consent to participate

The informed consent was waived with approval.

### Consent for publication

Not applicable.

### Code availability (software application or custom code)

Not applicable.

### Authors’ contributions

All authors contributed to the study conception and design. CJ and JY contributed to the literature search. YW, HFZ, TL, ZL, ZJJ, RQL, ZKW, FL, JZ, SBC, YL, HL, ZYL, YKL, HPZ, XBW and ZQR contributed to data collection and analysis. CJ, YW and JY contributed to data interpretation. CJ, CCW and JY contributed to writing of the manuscript.

## Acknowledgments

The authors thank all the participants for their contribution in this study.

